# Clinical Features and Outcomes of COVID-19 at a Teaching Hospital in Kingston, Jamaica

**DOI:** 10.1101/2022.04.27.22274270

**Authors:** Tamara Thompson, Yvonne Dawkins, Swane Rowe-Gardener, Lisa Chin-Harty, Kyaw Kyaw Hoe, Trevor S. Ferguson, Kelvin Ehikhametalor, Kelly Ann Gordon-Johnson, Varough Deyde

**Author notes:** Corresponding author -Tamara Thompson, Department of Medicine, University of The West Indies, Mona Campus, Kingston, Jamaica., Tele.

## Abstract

**Objective:** We examined the demographic, clinical characteristics and indicators of poor outcomes among hospitalized adults with COVID-19 at the University Hospital of the West Indies, Jamaica.

**Methods:** A retrospective medical record review between March 10 and December 31, 2020 analyzed data for 362 participants.

**Results:** There were 218 males (mean age 59.5 years) and 144 females (mean age 55.7 years). Hypertension, diabetes mellitus, cardiovascular disease, obesity and chronic kidney disease were the most common comorbidities. Cough, shortness of breath, fever and malaise were the most common presenting complaints. Fifty-one percent of patients were moderately to severely ill on admission; 11% were critically ill; 18% were admitted to the Intensive Care Unit (ICU). Death occurred in 62 (17%) patients (95% CI 13.6-21.4%). Among obese participants, there were increased odds of developing respiratory failure (OR 5.2, p < 0.001), acute kidney injury (OR 4.7, p < 0.001), sepsis (OR 2.9, p =0.013) and the need for ICU care (OR 3.7, p < 0.001). Factors independently associated with increased odds of death were age (OR 1.03 per year, p = 0.013) and obesity (OR 2.26, p = 0.017). Mortality also correlated significantly with D-dimer > 1000 ng/ml (OR 2.78; p = 0.03), serum albumin < 40 g/L (OR 3.54; p = 0.03) and serum LDH > 485 U/L OR 1.92, p = 0.11).

**Conclusions:** Targeted interventions among these high-risk patient subgroups may reduce in-patient morbidity and mortality.

## Introduction

Jamaica, the largest English-speaking island of the Caribbean, with a population of 2.9 million, recorded its first case of Severe Acute Respiratory Syndrome Coronavirus 2 (SARS-CoV-2) infection on March 10, 2020, a day before the World Health Organization declared its associated coronavirus disease (COVID-19), a global pandemic (1). By December 31, 2020, there was a confirmed total of 12,915 cases of COVID-19 on the island and 303 deaths (2,3).

The clinical presentation of COVID-19 is heterogenous with early reports of cough, myalgia, loss of appetite, diarrhea, fever, headache, and asthenia, accounting for more than 45% of patients (4,5). Some studies have noted a racial difference in laboratory abnormalities among COVID-19 patients, with higher percentage of white patients having a low white-cell and lymphocyte counts when compared to Blacks, and a higher percentage of Black patients presenting with elevated levels of serum creatinine, aspartate transferase (AST), procalcitonin, and C-reactive protein when compared to whites (6).

The varied complications of COVID-19 are mainly a result of the generalized inflammatory state the virus induces. These complications include severe viral pneumonia with respiratory failure, multiorgan and systemic dysfunctions with sepsis, septic shock, and death (7,8). Several studies have described the correlation between COVID-19 disease severity and poor outcomes. Indicators of poor outcomes described include advancing age, male sex, pre-existing comorbidities, lymphopenia, decreased serum albumin and elevated inflammatory markers (9).

Having an under-resourced healthcare system, coupled with a largely under-vaccinated population, Jamaica has struggled with recurrent surges of COVID-19 cases. Descriptive studies of the clinical characteristics and outcomes of COVID-19 among Afro-Caribbean populations in the English-speaking Caribbean could not be traced in the medical literature. We describe results of a retrospective analysis of COVID-19 patients admitted to a tertiary teaching hospital in Kingston, Jamaica. These results can inform appropriate approaches to risk stratification and early treatment strategies in the management of COVID-19 with a view to mitigating poor outcomes.

## Methods

We conducted a retrospective case series study of adults with laboratory confirmed SARS-CoV-2 infection admitted to the University Hospital of the West Indies (UHWI), Kingston, Jamaica from March 10 to December 31, 2020. The primary aims were to describe, the demographic and clinical characteristics as well as risk factors associated with severe disease and mortality among hospitalized COVID-19 patients.

### Patient Selection and Procedure

Confirmed cases of COVID-19 were identified using the World Health Organization (WHO) case definition. WHO defines a confirmed case of COVID-19 as a person with laboratory confirmation of SARS-CoV-2 infection, irrespective of clinical signs and symptoms (10). Real-time polymerase chain reaction (PCR) was used to identify the SARS-CoV-2 Envelope and RNA-dependent RNA polymerase nucleic acid sequences based on the Pan American Health Organization (PAHO) molecular testing protocol (11).

All patients admitted to the UHWI COVID-19 isolation wards with PCR confirmed SARS-CoV-2 tests were identified through hospital admission and laboratory records. From these records, data related to participants’ demographics, vital statistics, laboratory results on admission to hospital, medical histories, clinical management and trajectory were abstracted.

Patients were included in the study if they were ≥ 18 years old, had laboratory confirmed SARS-CoV-2 infection, and admitted to UHWI between March 10 and December 31, 2020. Patients were excluded from the study if they were < 18 years old, had confirmed SARS-CoV-2 infection but did not require admission and patients with SARS-CoV-2 RT-PCR negative results. Ethical approval was obtained from the University of the West Indies Mona Campus Research Ethics Committee: **ECP 46 20/21** and the Centers for Disease Control and Prevention (CDC) before starting the study.

### Study Variables and Definitions

We retrospectively abstracted patient specific variables from the electronic patient records and laboratory information systems of the UHWI. These variables included demographic data such as age, sex, smoking history, pregnancy and comorbidities (diabetes, hypertension, asthma, chronic obstructive lung disease, malignancy, HIV, chronic kidney disease, and other chronic medical conditions). Patient symptoms included respiratory (cough, shortness of breath, chest pain, sore throat and sneezing), gastrointestinal (nausea and vomiting, diarrhea, abdominal pain) and non-specific symptoms such as loss of taste or smell, joint pain, headache and fever. COVID-19 severity of illness (mild, moderate, severe or critical disease) determination was as per the WHO severity of illness classification (12). Laboratory variables at the time of admission, included complete blood count, absolute lymphocyte count, electrolytes, liver function tests, D-dimer, lactate dehydrogenase and serum albumin.

Outcome variables were respiratory failure including acute respiratory distress syndrome (severe pneumonia with oxygenation impairment and ratio of arterial partial pressure of oxygen to fraction of inspired oxygen (PaO2/FiO2) < 300 mmHg), sepsis (known or suspected source of infection with associated systemic inflammatory response syndrome), secondary infections (concurrent infections such as bacteremia and superimposed bacterial pneumonia) and coagulopathy (concurrent excessive bleeding or recognized thrombosis). The outcome of acute kidney injury was based on the clinical assessment documented by the medical team managing the patient. Other clinical outcomes included admission to the intensive care unit and in-hospital mortality whilst in the COVID-19 unit.

### Statistical Analyses

Descriptive statistics were obtained for clinical and demographic characteristics using mean and standard deviation for continuous variables and number and percent for categorical variables. For variables with non-normal distribution, we report medians with 25^th^ and 75^th^ centiles. Sex-specific estimates and comparison of sex differences were computed using t-tests for continuous variables and chi-squared test or Fisher’s exact test for categorical variables as appropriate. We estimated in-patient mortality rates with proportions and 95% confidence intervals (CI) for the period. Bivariate analysis and multivariate logistic regression models were used to identify factors associated with in-patient mortality and severe disease. Multivariate logistic regression analyses were used to identify factors associated with in-patient mortality and disease severity. A p value <0.05 was considered statistically significant. All statistical analyses were done using Stata version 16 and/or IBM SPSS version 21.

## RESULTS

### Demographic characteristics

From March 10 to December 31, 2020, a total of 362 SAR-CoV-2 positive patients admitted to the UHWI were identified as being eligible for inclusion in the study. Two hundred and eighteen (60%) participants were males with a mean age of 59.5 (59.5±16.4) years. More males were represented in the 60-69 years age group (25%), compared to all other age groups. This contrasts to females, who with a mean age of 55.7 (55.7±17.1) years, were represented in the 50-59 years age group (27%) (Table 1).

**Table 1.** Baseline Characteristics of Patients Hospitalized with COVID-19.

| **Demographic characteristics** | **Female**<br>**n = 144 (%)** | **Male**<br>**n = 218 (%)** | **Total**<br>**N = 362 (%)** | **p-value** |
| --- | --- | --- | --- | --- |
| **Age (Mean ± SD) <sup>1</sup>** | 55.7 ± 17.1 | 59.5 ± 16.4 | 58.0 ± 16.7 | 0.983 |
| **Age category** |  |  |  |  |
| <30 | 10 (6.9) | 12 (5.5) | 22 (6) |  |
| 30-39 | 22 (15.2) | 16 (7.3) | 38 (10.5) |  |
| 40-49 | 16 (11.1) | 34 (15.6) | 50 (13.8) |  |
| 50-59 | 39 (27.0) | 35 (16) | 74 (20.4) |  |
| 60-69 | 23 (15.9) | 55 (25.2) | 78 (21.5) |  |
| 70-79 | 24 (16.6) | 41 (18.8) | 65 (17.9) |  |
| $\geq 80$ | 10 (6.9) | 25 (11.4) | 35 (9.6) | **0.008** |
| **Presenting symptom** |  |  |  |  |
| *Shortness of breath* | 84 (58.3) | 129 (59.1) | 213 (58.8) | 0.874 |
| *Cough* | 91 (63.1) | 139 (63.7) | 230 (63.5) | 0.913 |
| *Fever* | 66 (45.8) | 120 (55.0) | 186 (51.3) | 0.086 |
| *Anorexia* | 36 (25.0) | 71 (32.5) | 107 (29.5) | 0.122 |
| *Malaise* | 34 (23.6) | 67 (30.7) | 101 (27.9) | 0.139 |
| *Diarrhea* | 35 (24.3) | 43 (19.7) | 78 (21.5) | 0.299 |
| *Nausea & vomiting* | 30 (20.8) | 31 (14.2) | 61 (16.8) | 0.100 |
| *Chest pain* | 26 (18.0) | 32 (14.6) | 58 (16.0) | 0.391 |
| *Headache* | 18 (12.5) | 23 (10.5) | 41 (11.3) | 0.567 |
| *Joint pain* | 13 (9.0) | 16 (7.3) | 29 (8.0) | 0.562 |
| *Abdominal pain* | 10 (6.9) | 14 (6.4) | 24 (6.6) | 0.845 |
| *Sore throat* | 8 (5.5) | 13 (5.9) | 21 (5.8) | 0.871 |
| *Runny nose* | 7 (4.8) | 11 (5.0) | 18 (4.9) | 0.937 |
| *Loss of taste* | 6 (4.1) | 12 (5.5) | 18 (4.9) | 0.567 |
| *Loss of smell* | 6 (4.1) | 7 (3.2) | 13 (3.5) | 0.632 |
| *Sneezing* | 1 (0.6) | 0 (0) | 1 (0.2) | 0.218 |
| **Co-morbidities** |  |  |  |  |
| *Hypertension* | 86 (59.7) | 114 (52.2) | 200 (55.2) | 0.164 |
| *Diabetes* | 64 (44.4) | 86 (39.4) | 150 (41.4) | 0.345 |
| *Cardiovascular* | 32 (22.2) | 39 (17.8) | 71 (19.6) | 0.310 |
| *Obesity* | 31 (21.5) | 34 (15.6) | 65 (17.9) | 0.150 |
| *Chronic kidney disease* | 12 (12) | 22 (16.4) | 34 (14.5) | 0.343 |
| *Asthma* | 16 (11.1) | 20 (9.1) | 36 (9.9) | 0.547 |
| *Malignancy* | 11 (7.6) | 22 (10.0) | 33 (9.1) | 0.427 |
| *COPD* | 0 (0) | 5 (2.2) | 5 (1.3) | 0.067 |
| **Smoking history**<br>**(N=276)** |  |  |  |  |
| *Non-smoker* | 97 (35.1) | 119 (43.2) | 216 (78.3) | **0.001** |
| *Current smoker* | 4 (1.4) | 17 (6.6) | 21 (7.6) | **0.001** |
| *Ex-smoker* | 6 (2.2) | 33 (11.9) | 39 (14.1) | **0.001** |
| **Severity of illness at admission** |  |  |  |  |
| *Asymptomatic* | 29 (20.1) | 37 (16.9) | 66 (18.2) |  |
| *Mild* | 20 (13.8) | 51 (23.3) | 71 (19.6) |  |
| *Moderate* | 35 (24.3) | 57 (26.1) | 92 (25.4) |  |
| *Severe* | 42 (29.1) | 51 (23.3) | 93 (25.6) |  |
| *Critical* | 18 (12.5) | 22 (10.0) | 40 (11.0) |  |
| **ICU Admission** | 25 (17.3) | 40 (18.3) | 65 (17.9) | 0.811 |
| **Overall outcome** |  |  |  |  |
| *Alive* | 123 (85.4) | 177 (81.1) | 300 (82.8) |  |
| *Death*<sup>2</sup> | 21 (14.5) | 41 (18.8) | 62 (17.1) | 0.292 |

*COPD: Chronic Obstructive Pulmonary Disease*
*Percentages (%) for Female, Male, Total categories calculated as number of participants in each category / n (sample size) or N (study population) x 100*
*95% Confidence interval: <sup>1</sup> 56.3, 59.8; <sup>2</sup> 0.69,2.26*
*Those with significant p values are indicated in bold with p values < 0.05 indicating that the difference was statistically significant.*

Among enrolled participants, pre-existing hypertension (55%), diabetes mellitus (41%) cardiovascular disease (20%), obesity (18%) and chronic kidney disease (15%) were the most commonly reported comorbid illnesses (Table 1). These comorbid illnesses were observed primarily in men (Supplemental Figure 1), but the difference was not statistically significant.

Underlying respiratory illnesses were relatively uncommon. A smoking history was obtained for 276 participants, of whom non-smokers accounted for 216 (78%). Those who were identified as current smokers were few (7.6%) and 14% reported a previous smoking history (Table 1).

### Clinical Characteristics

#### Presenting Symptoms

Among all enrolled participants, cough (64%), shortness of breath (59%) and subjective fever (51%), were the most common symptoms reported, followed by malaise (28%) and gastrointestinal symptoms such as anorexia (30%), diarrhea (22%), nausea and vomiting (17%) and abdominal pain (6.6%). Runny nose (4.9%), loss of taste (4.9%), loss of smell (3.5%) and sneezing (0.2%) were uncommon (Table 1).

### Laboratory results

On admission, the median serum inflammatory markers for participants were above normal range, such that the median values for D-dimer (163 tests) was 1466 U/L and serum lactate dehydrogenase (LDH) (159 tests) was 485 ng/ml. Median values for bilirubin, alanine transferase, alkaline phosphatase were within range. Whilst the median serum aspartate transferase and gamma glutamyl transferase were elevated. Sex differences were observed for some laboratory tests. Males had lower median absolute lymphocyte count (ALC), serum albumin, serum LDH and D-dimer levels compared to their female counterparts (Table 2).

**Table 2.** Median Laboratory Values at Admission for Hospitalized Male and Female Covid-19 Patients.

| **Lab value on admission** | **Number providing data n (%)** | **Males Median (p25, p75)** | **Female Median (p25, p75)** | **Both sexes Median (p25, p75)** |
| --- | --- | --- | --- | --- |
| WBC x 10<sup>9</sup> per liter | 354 (98.0) | 10.7<br>(7.04, 14.2) | 10.9<br>(8.91, 12.7) | 10.7<br>(7.89, 13.4) |
| Absolute lymphocyte count x 10<sup>9</sup> cells/liter | 283 (78.2) | 1.09<br>(0.75, 1.53) | 1.43<br>(0.99, 2) | 1.18<br>(0.81, 1.72) |
| Platelet x 10<sup>9</sup> per liter | 351 (97.0) | 187<br>(147, 235) | 232<br>(178.5, 306.5) | 202<br>(155, 260) |
| Hemoglobin g/dl | 291 (80.4) | 13<br>(10.5, 14.7) | 11.5<br>(10, 12.7) | 12.2<br>(10.2, 14) |
| Serum creatinine μmol/L | 337 (93.1) | 103<br>(80, 142) | 73<br>(56, 100) | 90<br>(68, 129) |
| D-dimer U/L | 163 (45.0) | 1417<br>(568, 6023) | 1514<br>(524, 3974.5) | 1466<br>(530, 5229) |
| LDH ng/ml | 159 (43.9) | 479<br>(347, 683) | 489<br>(359, 722) | 485<br>(348, 705) |
| Total bilirubin μmol/L | 204 (36.3) | 8<br>(6, 12) | 7<br>(5, 12.6) | 7.45<br>(5, 12) |
| Direct bilirubin μmol/L | 200 (55.2) | 4<br>(3, 6) | 3<br>(2, 5) | 3<br>(2, 5) |
| Albumin g/L | 210 (58.0) | 33<br>(29, 37) | 35<br>(32, 40) | 34<br>(30, 39) |
| Aspartate transferase U/L | 203 (56.0) | 47<br>(35, 77) | 37<br>(26, 61) | 46<br>(30, 67) |
| Alanine transferase U/L | 127 (35.1) | 40<br>(24, 74) | 25<br>(15, 50.5) | 34<br>(20, 67) |
| Gamma glutamyl transferase U/L | 170 (47.0) | 79<br>(42, 142) | 72<br>(36.5, 153) | 75<br>(38, 144) |
| Alkaline phosphatase U/L | 154 (42.5) | 76<br>(59, 114.5) | 87<br>(66, 131) | 82<br>(63, 117) |

*Overall number of participants = 362. Number of Females = 144, Males = 218*
*p25 is the 25<sup>th</sup> centile, p75 is the 75<sup>th</sup> centile*
*WBC: White Blood Cell, LDH: Lactate Dehydrogenase*

### Treatment

A range of treatments were prescribed to study participants but 9.9% (326) of participants did not receive any treatment as they had non-severe disease. Of this group, 279 (77%), received empiric antibiotics, 267 (74%) were given anticoagulation and 206 (57%) received dexamethasone or equivalent steroids. Less commonly prescribed treatments were remdesivir (12%), hydroxychloroquine (2.2%), convalescent plasma (2.2%) and tocilizumab (1.6%). There was no statistical difference between the sexes who received any of these treatments (Supplemental Table 1). Supplemental oxygen therapy was commonly used with 248 (69%) participants receiving low flow oxygen (providing flow rates up to 15L/min) and 84 (32%) receiving high flow oxygen therapy (providing flow rates up to 60 L/min).

### Disease severity

Among all participants, 18% were asymptomatic and 20% had mild disease, with both sets being admitted for medical diseases other than COVID-19. Most patients were found to have moderate (25%) or severe disease (26%) while 11% were critically ill at admission. Eighteen percent of the patients were admitted to the ICU (Table 1).

There was a male predominance for all categories of disease severity, including ICU admissions (62%) (Table 1). Among moderately to critically ill disease categories, the most represented ages were the 60-69 age group (22%) followed by those 50-59 years (20%); only 11% were below the age of 40 years (Supplemental Figure 2).

### Risk Factors associated with Disease severity

#### Outcomes

Prevalence of adverse outcomes associated with COVID-19 were acute respiratory failure including acute respiratory distress syndrome (ARDS) (26%), acute kidney injury (AKI) (16%), ICU admission (18%) and death (17%). Sepsis, and coagulopathy were uncommon outcomes (Table 3).

**Table 3.** Relationship Between Clinical Outcomes and Sex Among Hospitalized COVID-19 Patients.

| **Outcome** | **Overall %**<br>**(95% CI)** | **Male %**<br>**(95% CI)** | **Female %**<br>**(95% CI)** | **Odds ratio**<br>**(M vs F)** | **P-value M vs F** |
| --- | --- | --- | --- | --- | --- |
| *Coagulopathy* | 4.1<br>(2.5, 6.8) | 3.2<br>(1.5, 6.6) | 5.6<br>(2.8, 10.7) | 0.56 | 0.279 |
| *Sepsis* | 8.8<br>(6.3, 12.2) | 8.3<br>(5.3, 12.7) | 9.7<br>(5.8, 15.7) | 0.84 | 0.631 |
| *AKI* | 16.0<br>(12.6, 20.2) | 19.7<br>(14.9, 25.6) | 10.4<br>(6.4, 16.6) | 2.11 | **0.020** |
| *Death* | 17.1<br>(13.6, 21.4) | 18.8<br>(14.1, 24.6) | 14.6<br>(9.7, 21.4) | 1.36 | 0.298 |
| *ICU admission* | 17.9<br>(14.3,22.2)) | 18.4<br>(13.7, 24.1) | 17.4<br>(11.9, 24.5) | 1.06 | 0.81 |
| *Respiratory Failure or ARDS* | 26.4<br>(22, 31) | 26.6<br>(21.2, 32.9) | 25.7<br>(19.2, 33.5) | 1.05 | 0.847 |

*AKI: Acute Kidney Injury, ARDS: Acute Respiratory Distress Syndrome, ICU: Intensive Care*
*Those with significant p-values are indicated in bold with p-values < 0.05 indicating that the difference was statistically significant.*

There was no significant association between sex and mortality (OR 1.3, p=0.298) (Table 3). There was a significant association between age category and mortality (p=0.31) Table 4. The likelihood of death among those 60-69 and 70-79 years was roughly 2-fold but did not reach statistical significance. Participants aged ≥ 80 years were almost 6 times more likely to die (p = 0.030) compared to patients in the age 30-39-year category (Table 4).

**Table 4.** Mortality Outcomes by Age Category Among Hospitalized COVID-19 Patients.

| Age Category* | Death n (%) | Odds ratio | 95% CI | P value |
| --- | --- | --- | --- | --- |
| <30 | 2 (9.0) | 1.00 | - | - |
| 30-39 | 3 (7.8) | 0.85 | 0.13,5.57 | 0.872 |
| 40-49 | 10 (20) | 2.5 | 0.49,12.51 | 0.265 |
| 50-59 | 11 (14.8) | 1.74 | 0.35,8.54 | 0.492 |
| 60-69 | 12 (15.3) | 1.81 | 0.37,8.81 | 0.458 |
| 70-79 | 11 (16.9) | 2.03 | 0.41,10.00 | 0.381 |
| ≥80 | 13 (37.1) | 5.9 | 1.18,29.47 | **0.030** |

\* $p=0.031$ i.e. chi squared test for association between age-category and death
Those with significant $p$ values are indicated in bold with $p$ values $< 0.05$ indicating that the difference was statistically significant.

Obesity was associated with increased odds of death (OR 2.3, p < 0.05) (Table 6, Figure 1). This significant association was not observed with other pre-existing comorbidities such as diabetes mellitus, hypertension, chronic obstructive pulmonary disease, and cardiovascular disease.

**Figure 1.**
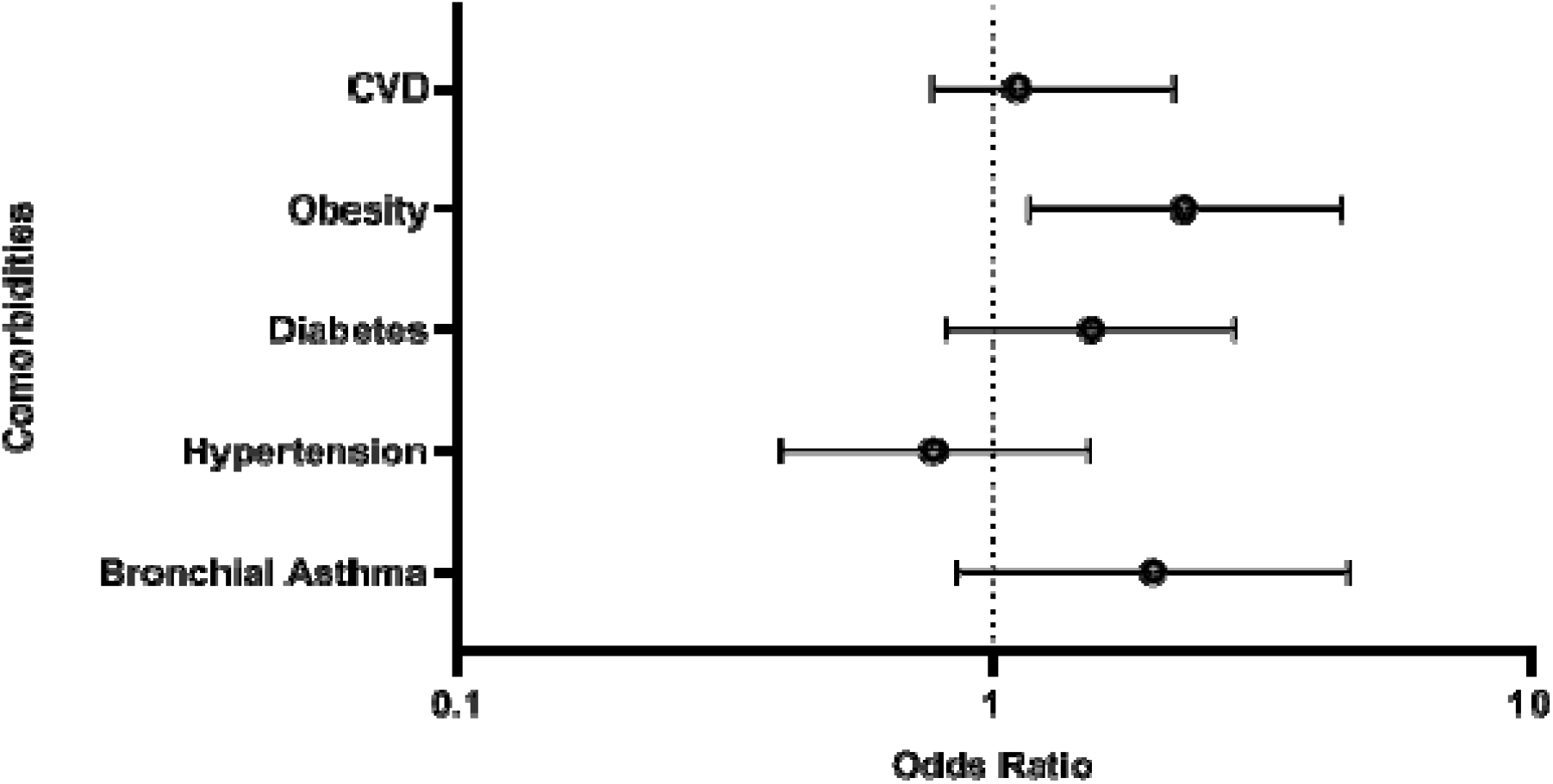
Age and Sex Adjusted Odds Ratio of Death in the Presence of Certain Comorbidities among COVID-19 Patients *Odds ratio for comorbidities associated with in-hospital mortality among COVID-19 patients after adjusting for age and sex. Relative to other comorbidities, obesity was associated with the greatest risk of death*.

The development of sepsis, secondary infections, and coagulopathy as outcomes among hospitalized patients were not significantly associated with sex, age or pre-existing comorbidities. However, obesity was associated with increased odds of developing sepsis as a complication (OR 2.92 (CI 1.25, 6.79; p < 0.05) (Table 5).

**Table 5.**
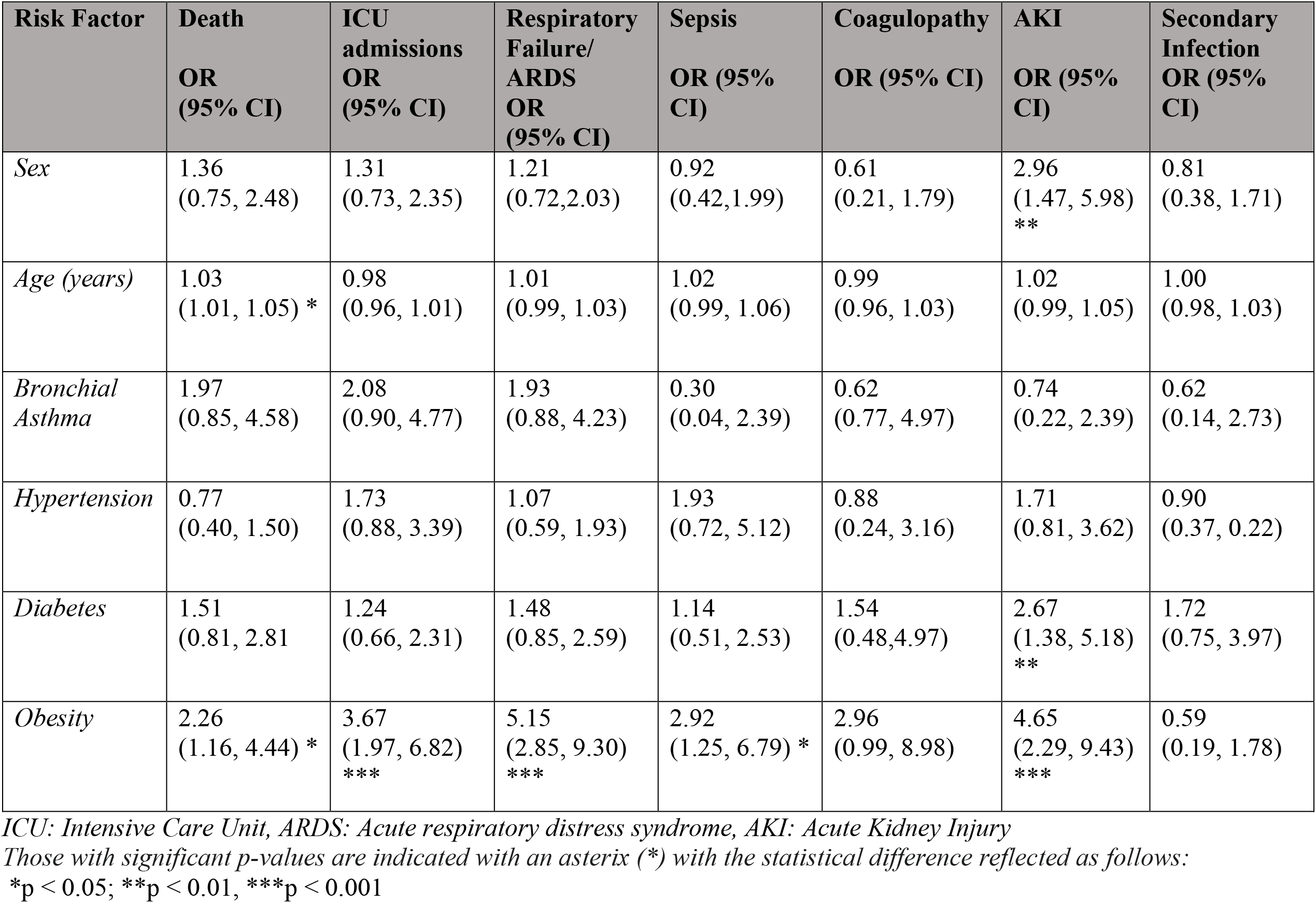
Multivariable Model of Association Between Clinical Outcomes with Age, Sex and Comorbidities.

Males had a 3-fold likelihood of developing AKI (p < 0.001). AKI was also significantly associated with pre-existing diabetes mellitus (OR 2.7, p < 0.01) and obesity (OR 4.7, p < 0.001). Additionally, among obese participants, the likelihood of developing respiratory failure including ARDS was 5-fold (p < 0.001) (Table 5).

There was an association between death and treatments. Of those who received high flow oxygen support, 42% died (OR 6.6, CI 3.68, 11.95, p < 0.001). Invasive mechanical ventilation was required in 41 (11%) patients with a mortality rate of 56% (OR 9.2, CI 3.72, 16.73, p < 0.001) (Supplemental Table2).

### Laboratory values at Admission and outcomes

Mortality was significantly higher in patients who had D-dimer > 1000 ng/ml [19 participants (20%); OR 2.74; p =0.04] and serum albumin < 40 g/L [36 participants (21.4%); OR 3.55; p = 0.04] than those who did not (Table 6). This was also observed in the multivariable analysis (Table 7). Serum LDH above the mean of 485 U/L was associated with 2-fold odds of death, but this was not statistically significant. There was also an increased odds of developing an ALC <1.0 × 10^9^ cells/liter (OR 2.24, p < 0.05), a WBC < 4 (OR 2.30, p < 0.05) or WBC > 11 × 10^9^ per liter (OR 2.14, p < 0.05) with the presence of obesity (Table 7). This finding was not associated with any other comorbidities.

**Table 6.** Association Between Mortality and Laboratory Variables.

| **Laboratory variable** | **Survived**<br>**N (%)** | **Died**<br>**N (%)** | **OR** | **95% CI** | **P value** |
| --- | --- | --- | --- | --- | --- |
| *D-dimer >1000*<br>*ng/ml* | 6 (8.7) | 19 (20.4) | 2.74 | 1.03, 7.27 | **0.043** |
| *serum albumin < 40*<br>*g/L* | 3 (7.1) | 36 (21.4) | 3.55 | 1.04, 12.14 | **0.044** |
| *ALC < 1.0 x*<br>*10<sup>9</sup> cells/liter* | 28 (15.4) | 24 (23.8) | 1.71 | 0.93, 3.15 | 0.083 |
| *LDH > 485 U/L* | 12 (15.0) | 20 (25.3) | 1.92 | 0.87, 4.26 | 0.108 |
| *WBC > 12 x 10<sup>9</sup> per*<br>*liter* | 32 (17.1) | 28 (16.8) | 0.98 | 0.56, 1.70 | 0.931 |

*ALC: Absolute Lymphocyte count, LDH: Lactate Dehydrogenase, WBC: White Blood Cell*
*N: number of patients with abnormal laboratory values as described on admission to hospital*
*n: number of deaths among patients with abnormal laboratory values on admission*
*Percentages (%): calculated based on $\frac{n}{N} \times 100$*
*p-values are from the logistic regression model for association between laboratory characteristic and death. Those with significant p-values are indicated in bold with p-values < 0.05 indicating that the difference was statistically significant.*

**Table 7.** Multivariable Model for Laboratory Characteristics and Association with Death Adjusted for Age, Sex and Comorbidities.

| **Risk Factors** | **D-Dimer >1000 ng/ml** | **Albumin <40 g/L** | **LDH >485 U/L** | **ALC <1.0 x 10<sup>9</sup> cells/liter** | **WBC <4 x 10<sup>9</sup> per liter** | **WBC >11 x 10<sup>9</sup> per liter** |
| --- | --- | --- | --- | --- | --- | --- |
| *Odds for Death* | 2.85 (0.98, 8.31) | 3.16 (0.87, 11.4) | 2.01 (0.88, 4.60) | 1.58 (0.83, 3.00) | 1.34 (0.33, 5.50) | 0.94 (0.52, 1.68) |
| *Age (years)* | 1.02 (0.99, 1.06) | 1.02 (0.99, 1.05) | 1.01 (0.98, 1.04) | 1.02 (0.99, 1.04) | 1.03 (1.01, 1.05) * | 1.02 (1.00, 1.04) * |
| *Sex* | 1.15 (0.44, 3.03) | 0.83 ((0.39, 1.78) | 1.06 (0.46, 2.44) | 1.44 (0.72, 2.85) | 1.35 (0.74, 2.46) | 1.41 (0.77, 2.06) |
| *Bronchial Asthma* | 4.25 (1.12, 15.5) | 2.14 (0.73, 6.24) | 2.74 (0.87, 8.59) | 1.62 (0.59, 4.50) | 1.90 (0.80, 4.50) | 1.80 (0.75, 4.32) |
| *Hypertension* | 0.85 (0.30, 2.44) | 0.71 (0.29, 1.66) | 0.70 (0.26, 1.88) | 0.66 (0.32, 1.36) | 0.77 (0.40, 1.50) | 0.85 (0.44, 1.67) |
| *Diabetes* | 1.68 (0.63, 4.47) | 1.26 (0.57, 2.77) | 2.00 (0.86, 4.81) | 1.74 (0.88, 3.44) | 1.50 (0.80, 2.79) | 1.53 (0.82, 2.88) |
| *Obesity* | 2.55 (0.99, 6.59) | 1.91 (0.85, 4.30) | 1.38 (0.56, 3.45) | 2.24 (1.08, 4.48) * | 2.30 (1.17, 2.79) * | 2.14 (1.06, 4.32) * |

*ALC = Absolute Lymphocyte count, LDH = Lactate Dehydrogenase, WBC = White Blood Cell Those with significant p-values (p<0.05) are indicated with an asterix (\*)*

## DISCUSSION

This single-center retrospective study is the first to summarize the demographic, clinical characteristics, and outcomes among adult hospitalized COVID-19 patients in Jamaica, the largest English-Speaking Caribbean island. These hospitalizations were in the first year of the global pandemic before the island-wide COVID-19 vaccination program had been instituted. The epidemic curve demonstrated a propagated spread of the infection after August 2020 following the phased re-opening of the Jamaican borders and the relaxation of some public health measures. During this time, the ancestral SARS-CoV-2 and later Alpha variants were in circulation.

The study found that there was a male predominance among admitted patients; the greatest burden of admissions was within the age 60-69 years group and the mean age was 58 years. Over half (55%) of patients had 1-2 comorbidities, but pre-existing respiratory diseases and current smoking history were uncommonly reported. Cough (64%) and shortness of breath (59%) were the most common presenting symptoms and fever was present in only 51% of patients. Most patients were identified on admission as being moderate to severely ill, and 11% were critically ill. The most common outcomes among patients were acute respiratory failure (26%), ICU admission (18%), AKI (16%), and death (17%). At admission, D-dimer > 1000 U/L, serum LDH > 485 ng/ml and serum albumin < 40 g/L were significantly associated with increased mortality. Additionally, patients over age 80 years had a 6-fold increased risk of dying, and obesity was independently associated with the risk of developing, respiratory failure, AKI sepsis and death.

The observation of an excess of hospitalized cases of COVID-19 among older adults, has been corroborated in other studies. One large retrospective USA analysis revealed a mean age of 61.2 (±17.9) years with younger ages of 41.7 (± 16.3) and 50.6 (range 16-94) years recorded among hospitalized patients in Bangladesh and Cuba (13–15). The disproportionate severity of COVID-19 among older age groups may be due in part to the increased number of comorbidities among the aged, and biological factors such as lower numbers of functional cilia, immunosenescence, inflamm-aging and lower levels of angiotensin converting enzyme 2 (ACE-2) (16,17).

In this study, a male preponderance was observed among patients with comorbidities, COVID-19 disease severity and mortality. Whilst a meta-analysis investigating sex differences among COVID-19 patients failed to show differences in SARS-CoV-2 infections between the sexes, it did demonstrate higher odds of ICU admissions and mortality among men (18). These findings were also noticed in previous human coronavirus outbreaks, involving SARS-CoV-1 and MERSCoV (19,20). Biological suggestions for these sex differences include the presence of X-linked genes which code for a stronger innate and adaptive immune system response. Additionally, the ACE-2 gene being X-linked, coupled with the upregulation of ACE-2 by estrogen results in less severe lung injury among females compared to males (21).

The top 4 reported comorbidities in this study were hypertension, diabetes, cardiovascular diseases, and obesity. These findings are consistent with that seen in other studies and reflect the chronic illnesses also seen in the wider Jamaican population (22–24).

Among obese participants, there were increased odds of developing respiratory failure (OR 5.2, p < 0.001), acute kidney injury (OR 4.7, p < 0.001), sepsis (OR 2.9, p =0.013) and the need for ICU care (OR 3.7, p < 0.001). It is known that obesity is associated with chronic inflammation, alteration in immune cell function, compromised T cell regulation and induction of a pro-inflammatory state of peripheral blood mononuclear cells (25–27). Additionally, decreased chest wall and lung compliance among obese patients, sets the stage for ventilatory compromise and death (28).

Cough, shortness of breath, fever and malaise ranked among the top reported symptoms among the study participants. Global data report fever and new cough as the most common COVID-19 symptoms. The presence of SARS-CoV-2 RNA has been detected in non-respiratory tissues via PCR. In these sites, tissue specific injury may occur in severe disease due in part to virus induce microangiopathy, direct viral cytopathic effects, and a dysregulated inflammatory response induced by injured endothelial cells and activated immune cells (29). This explains the range of non-respiratory pathologies seen among COVID-19 patients and their corresponding laboratory abnormalities. In this study, gastrointestinal symptoms were reported second to respiratory symptoms and AKI was a notable complication particularly among males, diabetic and older patients. With respect to laboratory derangements, lymphocyte depletion, elevations in D-dimer, abnormal liver biochemistries, elevated troponins, acute tubular injury, and elevations in acute phase reactants, have all been shown to be poor prognostic markers among COVID-19 patients.

Lymphocyte depletion with ALC < 1 cells/L has consistently been shown to be a marker of severe and critical COVID-19 (30). This study demonstrated 1.7-fold odds of death among participants with ALC < 1 cells/L, though this did not reach statistical significance. Other important biomarkers of COVID-19 disease severity include serum LDH, hypoalbuminemia and D-dimer. Elevated D-dimer, a marker of coagulation dysfunction, can predict critical illness and fatal outcomes among COVID-19 patients with a sensitivity of 75% and specificity of 83% (31,32). Hypoalbuminemia is an independent risk factor for death in COVID-19 (33,34). A pooled analysis of COVID-19 disease severity and mortality reported that elevated LDH was associated with a more than 6-fold increased odds of severe disease and a more than 16-fold odds of mortality (35). This study demonstrated that median D-dimer on admission was more than 2 times above the upper limit of normal and that a D-dimer >1000 ng/ml was a predictor of death. Further, serum albumin < 40 g/L and elevations of serum LDH above the mean of 485 U/L was associated with 2-fold odds of death. These findings corroborate reports from similar published studies.

### Limitations

This study is to be interpreted in the context of a number of limitations. This study was conducted during a time period when the ancestral and later Alpha SARS-CoV-2 variants were circulating in Jamaica. These findings therefore may not be representative of subsequent variants, particularly Delta and Omicron. Only patients diagnosed via SARS-CoV-2 PCR testing were included in the study. Excluded were hospitalized patients with a COVID-19-like illness who had a negative PCR, positive antigen or antibody test and any other adjunctive diagnostics consistent with COVID-19 such as specific radiographic findings. Not all patients had data for the laboratory investigations assessed or the full range of investigations, limiting a more fulsome review of all the laboratory data which could correlate with clinical outcomes. There was no consistent documentation of weight and height of study participants in the electronic patient records, hence documentation of obesity was based on the attending clinician’s subjective assessment of the patient’s body mass index. The study site is a major referral hospital in Jamaica, accepting severe and critically ill patients for specialist care. The population of COVID-19 patients sampled for this study therefore may include a bias towards the more severely and critically affected. Mortality thus could be excessive in this population. With limitations in ICU bed capacity, and the surge of patients on the hospitals, many critically ill COVID-19 patients were managed outside of the ICU setting. ICU admissions as an outcome measure for this study was thus grossly under-estimated. The study population included only Jamaican patients admitted to a single hospital site, thus affecting the generalizability of our results to larger populations affected by COVID-19.

## Conclusions

COVID-19 disease related morbidity continues to adversely impact health care systems. Insights into the specific clinical characteristics and outcomes among COVID-19 patients is important in identifying patients at highest risk for poor outcomes including death. Such patients can be quickly identified through risk stratification and distinguished from low-risk groups that may not need hospitalization. In this study we found that older age, male sex, and obesity correlate consistently with COVID-19 mortality and that elevated D-dimer, serum LDH and hypoalbuminemia are predictors of poor outcomes among hospitalized patients. Further studies on a larger Jamaican or Afro-Caribbean cohort are needed to corroborate these findings.

## Supporting information

ICMJE DISCLOSURE FORM

STROBE Statement Checklist

## Data Availability

All data produced in the present study are available upon request to the authors.

## Acknowledgments

The authors thank Heather Stewart for assistance in collating data for this manuscript. We also thank Anthony Myers for the development of the data abstraction tool and providing overall database management and technical support. The co-authors from the University of the West Indies, Faculty of Medical Sciences, acknowledge and are grateful for the support, oversight and guidance from the collaborators and co-authors from the Centers for Disease Control and Prevention on this study.

## Financial Support

The study was supported through the Centers for Disease Control and Prevention CDC#19JM3710P1001. The content of this manuscript is solely the responsibility of the authors and does not necessarily represent the official views of the Centers for Disease Control and Prevention.

## Disclosures

The authors have declared that no competing interests exist.

## Co-authors’ contact information

Yvonne Dawkins -

Swane Rowe-Gardener -

Lisa Chin-Harty -

Kyaw Kyaw Hoe -

Kelvin Ehikhametalor -

Trevor Ferguson -

Kelly-Ann Gordon-Johnson -

Varough Deyde -

## Notes

### Competing Interest Statement

The authors have declared no competing interest.

### Funding Statement

The study was supported through the Centers for Disease Control and Prevention CDC#19JM3710P1001. Authors from the CDC Caribbean Regional Office, Jamaica, are included on this study. No third party collaborated or provided funding to this project.The content of this manuscript is solely the responsibility of the authors and does not necessarily represent the official views of the Centers for Disease Control and Prevention.

### Author Declarations

Mona Campus Research Ethics Committee of University of the West Indies gave ethical approval for this work. Ethics committee of Centers for Disease Control and Prevention (CDC) gave ethical approval for this work.

