## Supplemental Figures 1 and 2; Supplemental Tables 1 and 2 for "Clinical Features and Outcomes of COVID-19 at a Teaching Hospital in Kingston, Jamaica"

**Supplementary Materials**

**Supplemental Figure 1 -** Frequency of Comorbidities by Sex (n=362)

Number of participants

| Number of pre-existing comordities | Males | Females | Total |
| --- | --- | --- | --- |
| 0 | 47 | 33 | 80 |
| 1 | 67 | 30 | 97 |
| 2 | 64 | 39 | 103 |
| 3 | 36 | 36 | 72 |
| 4 | 3 | 6 | 9 |
| 5 | 1 | 0 | 1 |

*The frequency distribution of comorbidities among hospitalized COVID-19 patients. As shown, most patients reported having 1 to 2 pre-existing comorbidities.*

**Supplemental Figure 2 -** Distribution of Moderate to Critically Ill Disease Severity by Age (N=225)

Number of participants

| Age category in years | Total |
| --- | --- |
| <30 | 6 |
| 30-39 | 19 |
| 40-49 | 35 |
| 50-59 | 50 |
| 60-69 | 51 |
| 70-79 | 42 |
| >80 | 22 |

*Distribution by age intervals of all adult patients with moderate to critically ill COVID-19. As shown, those patients most affected were between the ages of 50 and 69 years.*

**Supplemental Table 1. Prescribed Treatment for Hospitalized COVID-19 Patients by Sex**

| **Treatment given** | **Female**  **n = 144 (%)** | **Male**  **n = 218 (%)** | **Total**  **N = 362 (%)** | **P value** |
| --- | --- | --- | --- | --- |
| *Empiric antibiotics* | 108 (75) | 171 (78.4) | 279 (77.0) | 0.446 |
| *Anticoagulation* | 106 (73.6) | 161 (73.8) | 267 (73.7) | 0.959 |
| *Low flow oxygen* | 95 (65.9) | 153 (70.1) | 248 (68.5) | 0.399 |
| *Dexamethasone* | 84 (58.3) | 122 (55.9) | 206 (56.9) | 0.656 |
| *High flow oxygen* | 32 (22.2) | 52 (23.6) | 84 (32.2) | 0.719 |
| *Remdesivir* | 18 (12.5) | 27 (12.3) | 45 (12.4) | 0.974 |
| *Invasive ventilation* | 17 (11.8) | 24 (11.0) | 41 (11.3) | 0.815 |
| *Haemodialysis* | 5 (3.4) | 22 (10.0) | 27 (7.4) | **0.019** |
| *Hydroxychloroquine* | 2 (1.3) | 6 (2.7) | 8 (2.2) | 0.485 |
| *Convalescent plasma* | 1 (0.6) | 7 (3.2) | 8 (2.2) | 0.153 |
| *Tocilizumab* | 1 (0.6) | 5 (2.2) | 6 (1.6) | 0.409 |
| *No treatment* | 17 (11.8) | 19 (8.7) | 36 (9.9) | 0.336 |

- *p-value from chi squared test for difference in proportion by sex; where there were cell values <5 we report p-values from Fisher’s exact test.*
- *Those with significant p values are indicated in bold with p values < 0.05 indicating that the difference was statistically significant*

**Supplemental Table 2. Correlation of Treatment and Mortality Among Hospitalized Covid-19 Patients**

| **Treatment given** | **Recipients n (%)** | **Deaths n (%)** | **OR** | **95% CI** | **p-value** |
| --- | --- | --- | --- | --- | --- |
| *Empiric Antibiotics* | 279 (77.0) | 51 (18.2) | 1.5 | 0.72, 2.95 | 0.288 |
| *Anticoagulation* | 267 (73.7) | 46 (17.2) | 1.02 | 0.55, 1.92 | 0.932 |
| *Low flow oxygen* | 248 (68.5) | 47 (18.9) | 1.5 | 0.82, 2.89 | 0.176 |
| *Dexamethasone* | 206 (56.9) | 36 (17.4) | 1.1 | 0.61, 1.84 | 0.840 |
| *High flow oxygen* | 84 (23.2) | 35 (41.6) | 6.6 | 3.68, 11.95 | **<0.001** |
| *Remdesivir* | 45 (12.4) | 14 (31.5) | 2.5 | 1.25, 5.1 | **0.010** |
| *Mechanical ventilation* | 41 (11.3) | 23 (56) | 9.2 | 3.72, 16.73 | **<0.001** |
| *Haemodialysis* | 27 (7.4) | 11 (40.7) | 3.8 | 1.68, 8.72 | **0.001** |
| *Hydroxychloroquine* | 8 (2.2) | 4 (50) | 5.1 | 1.24, 20.99 | **0.024** |
| *Convalescent Plasma* | 8 (2.2) | 5 (62.5) | 8.68 | 2.01, 37.36 | **0.004** |
| *Tocilizumab* | 6 (1.6) | 4 (66.6) | 10.2 | 1.83, 57.41 | **0.008** |

- *p-value from chi squared test for difference in proportion by sex; where there were cell values <5 we report p-values from Fisher’s exact test.*
- *Those with significant p values are indicated in bold with p values < 0.05 indicating that the difference was statistically significant*
